# Attribution of invasive group A streptococcal infections to varicella and respiratory virus circulation, the Netherlands, 2010-2023

**DOI:** 10.1101/2024.01.09.24300975

**Authors:** Brechje de Gier, Jan van de Kassteele, Liselotte van Asten, Annelot Schoffelen, ISIS-AR study group, Mariette Hooiveld, Margreet J.M. te Wierik, Nina M. van Sorge, Hester E. de Melker

## Abstract

**Background:** After the lifting of most COVID-19 control measures, many infectious diseases re-emerged in 2022. A strong increase in invasive group A streptococcal (iGAS) infections, among both adults and young children, was reported by several countries. Viral infections such as influenza and varicella, known risk factors for iGAS infection, also increased during 2022. We estimated the proportion of GAS skin and soft tissue infections (SSTI) in children aged 0-5 years attributable to varicella, and the proportion of GAS pneumonia/sepsis in children aged 0-5 and adults attributable to respiratory viruses in the Netherlands.

**Methods:** We performed time-series regression using weekly data on influenza A and B, RSV, hMPV and SARS-CoV-2, varicella and non-invasive GAS infections, and GAS isolates cultured from blood, lower airways, skin, pus and wounds, from January 2010 to March 2023.

**Results:** Up to 2022, approximately 50%(95%CI 36-64%) of GAS SSTI in children were attributable to varicella. Between January 2022 and March 2023, 34%(95%CI 24-43%) of GAS SSTI cases were attributable to varicella. Of iGAS pneumonia/sepsis between January 2022 and March 2023, 25%(95%CI 18-31%) and 37%(95% CI 23-53%) was attributable to respiratory virus infections in adults and children, respectively, with the largest contributor being influenza A.

**Conclusions:** Varicella and respiratory virus infections likely contributed to, but only partly explain, the observed iGAS increase among children and adults in 2022-2023 in the Netherlands. Public health measures to control viral infections, such as vaccination against varicella or influenza, might reduce the iGAS disease burden but will not suffice to curb the current high incidence.

## Background

In 2022-2023, an increase in the number of invasive group A streptococcal (iGAS) infections was reported by several countries, including the Netherlands [1, 2]. While many infectious diseases showed a strong re-emergence after lifting of COVID-19 control measures, the re-emergence of iGAS seemed disproportionate to a mere catch-up effect from 2020-2021. A strong iGAS surge was initially observed among young children, showing a sevenfold increase in iGAS infections among children aged 0-5 years in the first half of 2022 compared to 2016-2019. In particular, the high number of cases presenting with necrotizing fasciitis was unprecendented [1]. From November 2022 onward, a further surge in iGAS infections occurred, both in children and in adults, with pleural empyema and pneumonia being reported by clinicians to occur more than expected. This was also observed in the United Kingdom, France, Spain, and Portugal [3–6].

Specific viral infections increase the risk of iGAS infection. For children, varicella is a well-known risk factor for developing iGAS infection [7], as well as a more severe iGAS disease course [8]. Also, influenza infection is a known risk factor for iGAS infection [9, 10]. Influenza A and varicella, like iGAS, reached a high incidence in early 2022. A Dutch pediatric survey showed that varicella preceded or coincided with 33% of iGAS cases, and that 18% of iGAS infections were preceded by confirmed influenza infection [11]. Surveillance data from England showed respiratory syncytial virus (RSV) and human metapneumovirus (hMPV) to be the most common co-infecting viruses in pediatric GAS in October-November 2022 [3]. A Portuguese study on pediatric iGAS between September 2022 and May 2023 reported varicella and upper respiratory infection to each precede iGAS in 24.6% of cases [5]. As varicella is mostly associated with GAS skin and soft tissue infection (SSTI) and respiratory viruses are risk factors for GAS pneumonia or sepsis, we hypothesized that increased viral infections could explain the observed high incidence of GAS SSTI (for varicella) and GAS pneumonia and sepsis (for respiratory viruses)[4].

We used existing separate registries of the different infectious diseases and performed time-series regression analysis to assess the possible contribution of re-emergence of these viral infections on iGAS. Our study period includes January 2010 - March 2023. We aimed to quantify the association between varicella and GAS SSTI in children aged 0-5 years old, and the association between influenza A, influenza B, RSV, hMPV, and SARS-CoV-2 and GAS pneumonia or sepsis (PS) in children aged 0-5 years old and in adults. Our study quantifies temporal associations between numbers of iGAS infections and numbers of viral infections 0-4 weeks prior. If a causal link between these types of infections is assumed, these associations can be interpreted as attributable proportions.

## Methods

### Data sources

#### iGAS infections

We used the weekly number of GAS cultures extracted from the database of the Dutch Infectious Diseases Surveillance Information System – Antimicrobial Resistance (ISIS-AR), as a measure for the number of iGAS infections [12]. Based on the year and month of birth, weekly numbers of isolates were obtained for children aged 0-5 years old and for adults aged 18 and older. As a proxy for the number of GAS SSTIs, we extracted the number of GAS isolates cultured from skin, wound or pus. The numbers of GAS pneumonia/sepsis were approximated as the number of GAS isolates cultured from blood or the lower airway.

Laboratories participating in ISIS-AR provide data for all isolates from medical routine diagnostics for which antibiotic susceptibility tests were performed, regardless of the pathogen. The database contains data from all types of healthcare facilities, including hospitals, primary care and long-term care facilities. In case of multiple isolates per patient within 3 months, we included only the first isolate in our analysis based on the assumption that multiple isolates within 3 months stem from the same infection episode. The number of laboratories participating in ISIS-AR gradually increased during the study period, from an average of 32 laboratories per week in 2010 to 42 in 2014, after which the weekly number of participating laboratories stabilized (Figure S3). The total number of laboratories in the Netherlands decreased during the study period from approximately 60 in 2010 to 50 in 2023. Because the catchment population of participating laboratories was unknown, a true population coverage could not be obtained.

#### Non-invasive GAS infections

The Nivel primary care database is a surveillance system monitoring general practitioner (GP) visits for a number of infectious disease syndromes [13]. From this database, the weekly number of visits by children aged 0-4 with diagnosis streptococcal pharyngitis/scarlet fever (ICPC code R72, henceforth “non-invasive GAS infection”) per 100.000 population was obtained as a proxy for the incidence of non-invasive GAS infections in young children. This timeseries was used as a representation of the circulation of GAS in general.

#### Varicella zoster virus infections

The number of GP visits for varicella zoster (ICPC code A72) by children aged 0-4 was obtained per week from the Nivel primary care database.

#### Respiratory viruses

The weekly numbers of detections of influenza A, influenza B, RSV and hMPV were obtained from the Virological Weekly Report. This is a surveillance system collecting aggregated numbers of virus detections per laboratory per week, and does not contain any patient-level information.

#### SARS-CoV-2

The national average SARS-CoV-2 RNA load per 100.000 inhabitants from wastewater surveillance was used as a proxy for SARS-CoV-2 infections in the population [14]. Data is available from 30 March 2020, the first SARS-CoV-2 infection in the Netherlands was detected on February 27, 2020. The natural logarithm of the RNA load served as indicator of SARS-CoV-2 incidence. From the abovementioned data sources, a dataset was generated with weekly values of determinants and outcomes, ranging from week 1, 2010 (January 4, 2010) to week 13, 2023 (March 27, 2023).

#### Model assumptions and construction

The model was based on the following assumptions:

– We assumed the iGAS counts in week *t* followed a Negative Binomial distribution, with expected value *µ_iGAS,t_* and overdisperson parameter *φ*. The variance is given by *µ_iGAS,t_*(1 + *µ_iGAS,t_*/*φ*). The overdispersion parameter was treated as a nuisance parameter, and assumed to be constant over the study period.
– We assumed changes in iGAS count to be attributable not only to circulation of non-invasive GAS and viruses, but also to unmeasured factors such as changes in population immunity and in the virulence of circulating GAS types.To allow for such explanatory unmeasured factors, a large scale time trend was included. The large scale trend term was modelled by penalized B-splines [15], reparametrized using an orthogonal decomposition to increase computational efficiency [16]. To ensure the trend is always positive, the exponent was taken.
– We assumed that antibiotic susceptibility is always tested for GAS isolates, as is the standard of care.
– We further assumed that all laboratories had roughly equal probabilities of contributing GAS isolates to the data. This weekly expected value of iGAS count was multiplied with the known proportion of laboratories reporting to ISIS-AR that week relative to the maximum number in the dataset (45), because the lower the number of participating laboratories, the less iGAS cases are expected in the data. In other words, the weekly attributions were corrected for the time-varying completeness of the surveillance data.
– The pathogen-specific regression parameters were assumed to be constant during the study period. Two exceptions are made: 1) Because we worked with the natural logarithm of the RNA load as indicator of SARS-CoV-2 incidence, *β_SARS-CoV-2_* describes how much the expected iGAS count changes if the lag-weighted RNA load is a factor *e* larger. 2) The regression coefficient of influenza A, *β_infA_*, was allowed to vary between influenza seasons (piece-wise constant between July of one year to June next year), since it is known that the severity of influenza varies between seasons and in a previous study we found that the association between influenza A and iGAS also varies between seasons [10].
– We assumed the included weekly counts of the pathogens to be possibly associated with iGAS in the same week and the four following weeks.

The weekly expected value *µ_iGAS,t_* was written as the sum of a large scale time trend *µ_trend,t_* and pathogen specific contributions *µ_path,i,t_*, where *i* is the *i*-th pathogen. The weekly contribution of each pathogen to the expected iGAS counts is given by a linear function as the weighted sum of the reported pathogen incidence in current week *t* to four weeks back *t* - 4: *µ_path,i,t_* = Σ*_j= 0..4_ w_i,j_ β_path,i_ path_i,t-j_*. The weights *w_i,j_* were included to model possible lag-effects of pathogen *i* [17], and sum up to one. The weights are pathogen-specific and were assumed to be constant during the study period. Since we explicitly used the identity link function, the regression parameters *β_path,i_* describe how much the expected iGAS count changes if the lag-weighted incidence of pathogen *i* changes one unit. To ensure attributions are always positive, *β_path,i_* was forced to be greater than zero. We did not include any harmonic terms in the models. Instead, as a proxy for seasonality of GAS transmission, we included the number of consultations for non-invasive GAS syndromes.

All statistical analyses were carried in R [18]. Because of the restrictions on the parameters (both the regression coefficients and the weights), the models were formulated in the Bayesian framework using priors. The models were fitted in Stan [19]. The regression parameters were given half-Normal(0, 1) distributions, the weights were given a Dirichlet(0.2, 0.2, 0.2, 0.2, 0.2) distribution, and the overdispersion parameter *φ* was given an Exponential(0.1) distribution.

#### Data and code availability

Data and code are available from https://github.com/kassteele/Papers/DeGier_iGAS_attribution

## Results

Over the entire study period, 1,595 GAS SSTI cases among children age 0-5 years old were included. For GAS pneumonia/sepsis, 446 cases were included among children 0-5 years old, and 4,643 adult cases were included. The observed time series are shown in Figures S1-S2. All GAS timeseries show a winter seasonality, although this is less clear for pneumonia/sepsis in children due to small numbers. The numbers of GAS SSTI and pneumonia/sepsis as well as GP consultations for non-invasive GAS syndromes were decreased throughout 2020 and 2021. The numbers of GAS SSTI and pneumonia/sepsis in 2022 exceeded the peaks from seasons before 2020, in line with the trend seen in the national iGAS notifications [1]. However, non-invasive GAS GP consultations had returned to the typical levels observed before the pandemic. For most viruses, the typical seasonality was interrupted in 2020 and 2021 concurrent with COVID-19 public health and social control measures. RSV and hMPV started to re-emerge already in 2021, while influenza A re-emerged in 2022 and influenza B in 2023 (Figure S2). Varicella GP consultations surged in the first months of 2022, followed by a very low incidence at the start of 2023 (Figure S1).

### GAS SSTI and varicella – children

The estimated absolute number and proportion of GAS SSTI attributable to varicella and non-invasive GAS circulation are presented in Figure 1 and Table 1. From 2010 to 2021, the proportion of GAS SSTI estimated to be attributable to varicella fluctuated around 50%. Non-invasive GAS circulation was estimated to account for around 40% of the GAS SSTI incidence, with the remaining 10% not attributable to varicella nor non-invasive GAS; this remaining unexplained proportion of GAS SSTI is reflected in the large-scale time trend (‘trend’ parameter). In the first half of 2022, still around 50% of GAS SSTI was estimated to be attributable to varicella. From July 2022 to March 2023, the attributable fraction of varicella diminished rapidly, and the proportion of GAS SSTI not explained by varicella or non-invasive GAS (‘trend’ parameter) increased to 70%. Overall from January 2022 to March 2023, 34% (95% CI 24-43%) of GAS SSTI was attributable to varicella; a lower proportion than in previous years.

**Figure 1.**
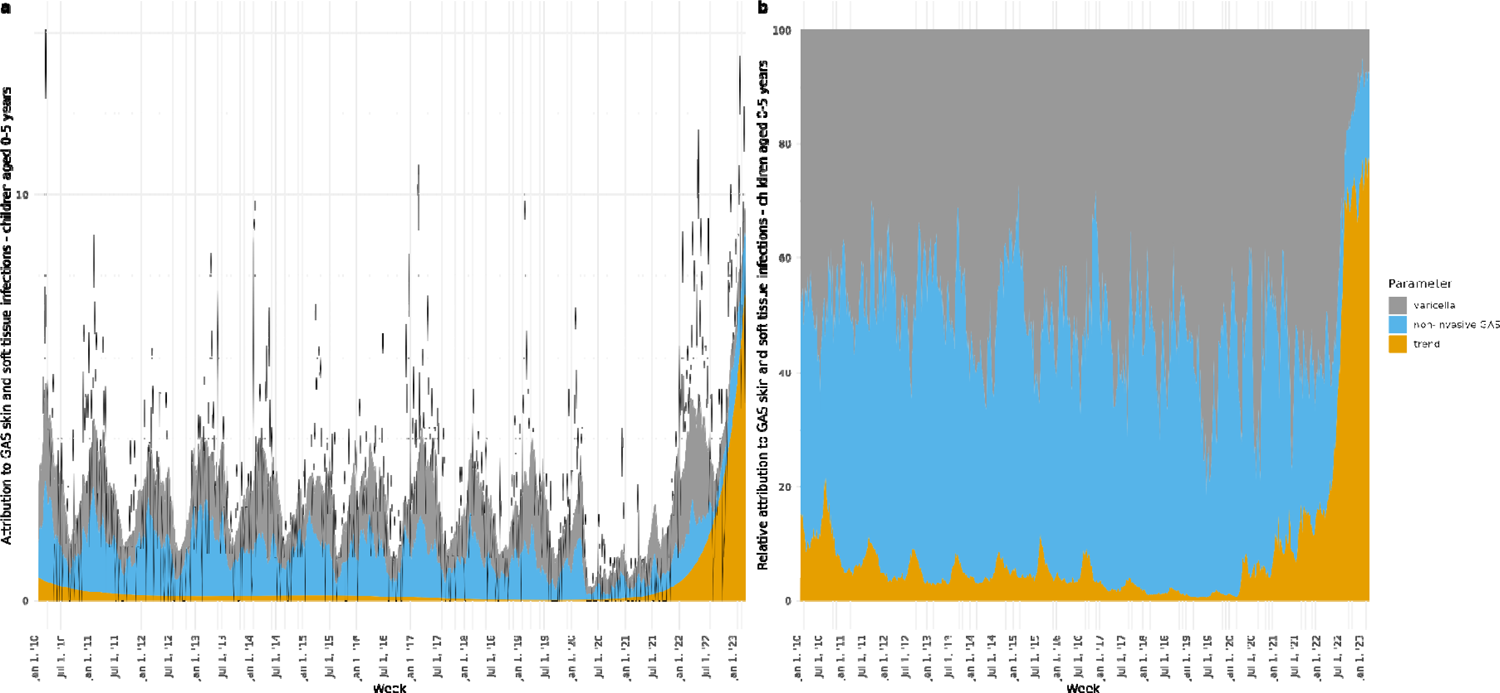
Estimated weekly absolute (A) and relative (B) attributions to parameters varicella, non-invasive GAS and long-term time trend to GAS skin and soft tissue infections in children aged 0-5 years, January 2010-March 2023. The black line in panel A shows the observed number of GAS skin and soft tissue infections.

**Table 1.**
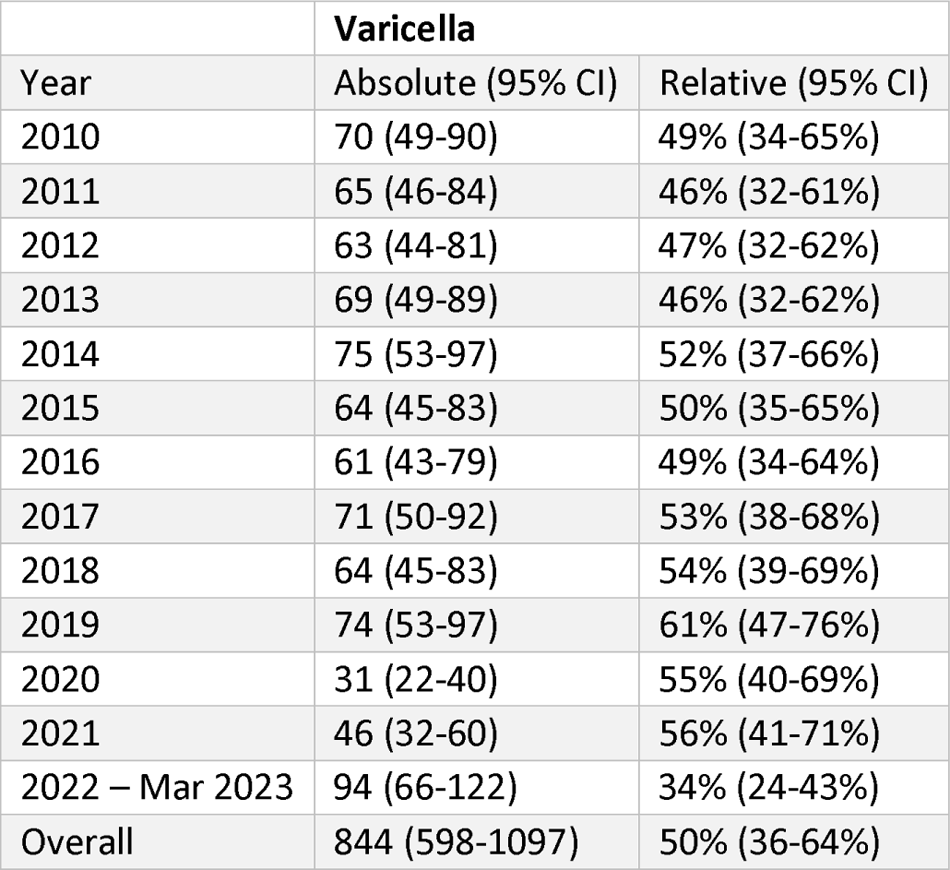
GAS skin and soft tissue infections in children 0-5 years old, attributed to varicella, January 2010-March 2023. Absolute numbers are summarized per year and the study period, while the relative attributions are averaged over years and the study period. CI: Confidence interval.

|  | **Varicella** |  |
| --- | --- | --- |
| Year | Absolute (95% CI) | Relative (95% CI) |
| 2010 | 70 (49-90) | 49% (34-65%) |
| 2011 | 65 (46-84) | 46% (32-61%) |
| 2012 | 63 (44-81) | 47% (32-62%) |
| 2013 | 69 (49-89) | 46% (32-62%) |
| 2014 | 75 (53-97) | 52% (37-66%) |
| 2015 | 64 (45-83) | 50% (35-65%) |
| 2016 | 61 (43-79) | 49% (34-64%) |
| 2017 | 71 (50-92) | 53% (38-68%) |
| 2018 | 64 (45-83) | 54% (39-69%) |
| 2019 | 74 (53-97) | 61% (47-76%) |
| 2020 | 31 (22-40) | 55% (40-69%) |
| 2021 | 46 (32-60) | 56% (41-71%) |
| 2022 – Mar 2023 | 94 (66-122) | 34% (24-43%) |
| Overall | 844 (598-1097) | 50% (36-64%) |

### GAS pneumonia/sepsis and respiratory virus infections – children

The estimated absolute and relative number of GAS pneumonia/sepsis in children aged 0-5 years attributable to respiratory viruses are presented in Figure 2 and Table 2. In our model, influenza A, RSV and hMPV all provided relevant contributions to GAS pneumonia/sepsis cases in children aged 0-5 years, accounting for high attributable fractions during the seasonal peaks of these viruses (Figure 4B). The largest attributable fraction over the entire study period was estimated for influenza A; 12% (95% CI 9-16%), ranging from 3% to 26%, depending on the year. Over the period January 2022-March 2022, the average attributable proportions were 17% (95%CI 9-28%) for influenza A, 11% (95%CI 2-21%) for RSV and 6% (95%CI 0-13%) for hMPV. SARS-CoV-2 did not seem to be a risk factor of concern for GAS pneumonia/sepsis incidence among children aged 0-5, with an estimated attributable fraction of only 3% in 2022-March 2023 (95% CI 0-10%), while this was a period with very high rates of SARS-CoV-2 infections. Despite high incidences of respiratory viruses, we estimated the number of GAS pneumonia/sepsis not attributable to respiratory viruses or non-invasive GAS circulation to have increased to 54% (95% CI 37-69%) in 2022-2023, compared to 40% (95% CI 21-61%) over the entire study period (Figure 4A, ‘trend’ parameter).

**Figure 2.**
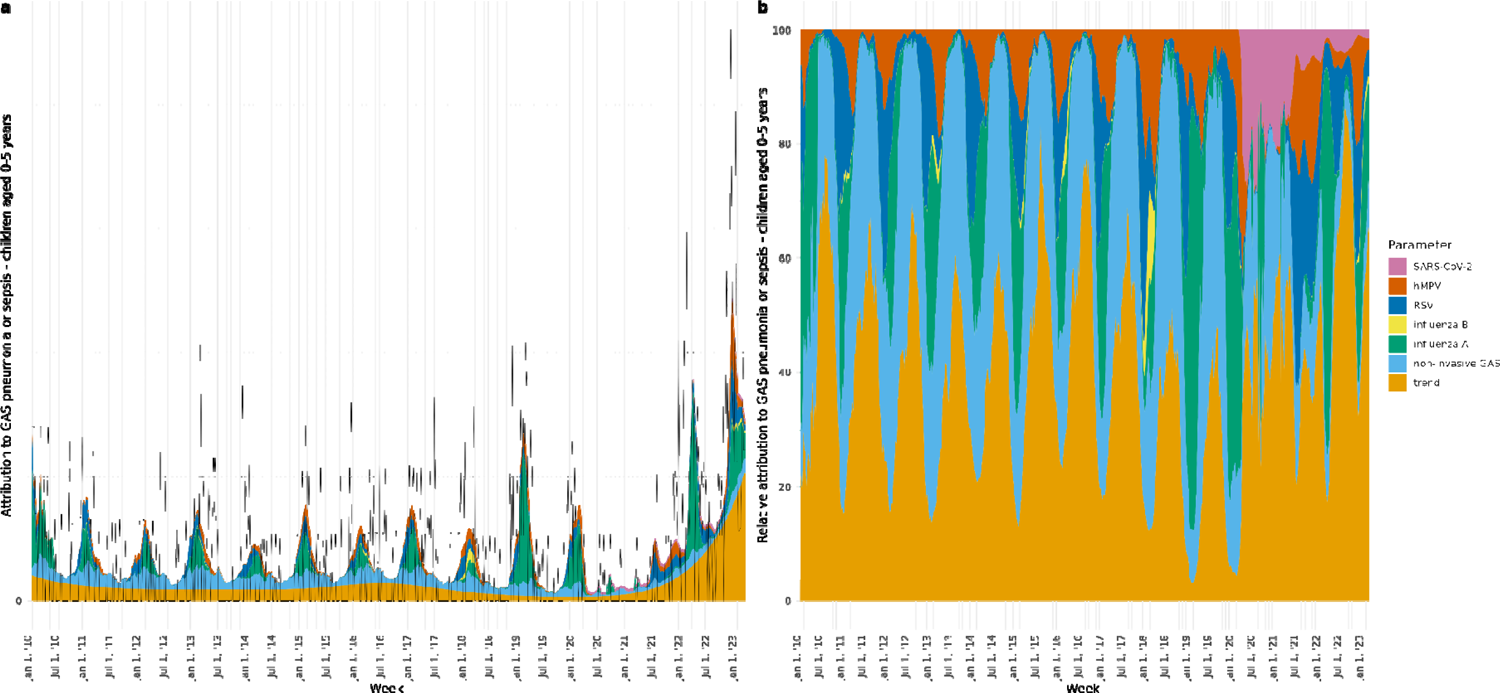
Estimated weekly absolute (A) and relative (B) attributions to parameters SARS-CoV-2, hMPV, RSV, influenza B, influenza A, non-invasive GAS and long-term time trend to GAS pneumonia or sepsis in children aged 0-5 years, January 2010-March 2023. The black line in panel A shows the observed number of GAS pneumonia or sepsis.

**Table 2.** GAS pneumonia or sepsis in children 0-5 years old, attributed to respiratory virus infections, January 2010-March 2023. Absolute numbers are summarized per year and the study period, while the relative attributions are averaged over years and the study period. *: the relative attributions to SARS-CoV-2 are averaged over April - December 2020 (for the year 2020) and over April 2020-March 2023 (overall). CI: Confidence interval.

|  | Influenza A |  | Influenza B |  | RSV |  | hMPV |  | SARS-CoV-2 |  | Respiratory viruses combined |  |
| --- | --- | --- | --- | --- | --- | --- | --- | --- | --- | --- | --- | --- |
| Year | Absolute (95% CI) | Relative (95% CI) | Absolute (95% CI) | Relative (95% CI) | Absolute (95% CI) | Relative (95% CI) | Absolute (95% CI) | Relative (95% CI) | Absolute (95% CI) | Relative * (95% CI) | Absolute (95% CI) | Relative (95% CI) |
| 2010 | 14 (2-30) | 19% (4-32%) | 0 (0-0) | 0% (0-0%) | 4 (1-7) | 7% (1-13%) | 2 (0-4) | 3% (0-6%) |  |  | 20 (7-36) | 29% (13-43%) |
| 2011 | 5 (0-15) | 7% (1-15%) | 0 (0-1) | 0% (0-1%) | 5 (1-9) | 10% (2-18%) | 2 (0-4) | 5% (0-12%) |  |  | 12 (5-22) | 22% (12-32%) |
| 2012 | 5 (0-13) | 9% (1-19%) | 0 (0-0) | 0% (0-0%) | 4 (1-8) | 10% (2-19%) | 2 (0-4) | 4% (0-9%) |  |  | 10 (4-20) | 23% (12-35%) |
| 2013 | 6 (0-16) | 10% (1-19%) | 0 (0-1) | 1% (0-2%) | 4 (1-7) | 8% (1-15%) | 2 (0-5) | 6% (0-13%) |  |  | 12 (5-23) | 24% (13-35%) |
| 2014 | 5 (1-13) | 11% (2-23%) | 0 (0-0) | 0% (0-0%) | 3 (0-5) | 8% (1-15%) | 2 (0-4) | 6% (0-13%) |  |  | 9 (4-17) | 24% (13-35%) |
| 2015 | 6 (1-15) | 9% (1-17%) | 0 (0-1) | 1% (0-2%) | 3 (1-7) | 7% (1-14%) | 3 (0-7) | 7% (0-15%) |  |  | 12 (6-22) | 23% (15-31%) |
| 2016 | 4 (1-11) | 7% (1-16%) | 0 (0-1) | 1% (0-3%) | 4 (1-7) | 8% (1-15%) | 2 (0-6) | 6% (0-13%) |  |  | 11 (5-18) | 22% (13-31%) |
| 2017 | 6 (0-16) | 9% (1-16%) | 0 (0-0) | 0% (0-1%) | 3 (0-6) | 7% (1-12%) | 3 (0-7) | 7% (0-16%) |  |  | 12 (6-22) | 23% (14-32%) |
| 2018 | 3 (1-8) | 7% (2-16%) | 2 (0-6) | 3% (0-10%) | 4 (1-8) | 10% (2-19%) | 4 (0-10) | 11% (1-23%) |  |  | 13 (7-21) | 31% (21-42%) |
| 2019 | 21 (10-35) | 26% (17-35%) | 0 (0-0) | 0% (0-0%) | 3 (1-7) | 8% (1-15%) | 4 (0-9) | 9% (1-21%) |  |  | 28 (17-42) | 43% (34-54%) |
| 2020 | 10 (3-21) | 21% (9-31%) | 0 (0-0) | 0% (0-0%) | 2 (0-4) | 5% (1-10%) | 3 (0-7) | 7% (1-15%) | 2 (0-5) | 20% (1-49%) | 17 (9-27) | 47% (30-69%) |
| 2021 | 1 (0-2) | 3% (1-5%) | 0 (0-0) | 0% (0-0%) | 6 (1-12) | 15% (3-29%) | 4 (0-9) | 10% (1-22%) | 2 (0-8) | 11% (0-32%) | 13 (6-20) | 39% (21-60%) |
| 2022- Mar 2023 | 33 (12-61) | 17% (9-28%) | 1 (0-3) | 0% (0-1%) | 15 (2-29) | 11% (2-21%) | 10 (1-23) | 6% (0-13%) | 3 (0-10) | 3% (0-10%) | 61 (36-90) | 37% (23-53%) |
| Overall | 119 (80-167) | 12% (9-16%) | 4 (0-14) | 0% (0-2%) | 59 (10-117) | 9% (2-16%) | 42 (3-98) | 7% (0-14%) | 7 (0-23) | 10% (0-27%) | 231 (170-297) | 30% (22-39%) |

### GAS pneumonia/sepsis and respiratory virus infections – adults

The estimated absolute number and proportion of GAS pneumonia/sepsis among adults attributed to respiratory viruses are presented in Figure 3 and Table 3. Overall, the proportion attributable to the combined included respiratory viral infections is lower in adults (13%, 95% CI 10-16%) than in children aged 0-5 years (30%, 95%CI 22-39%). Of the included respiratory viruses, influenza A accounted for the largest attributable fraction of GAS pneumonia/sepsis in adults with 8% (95% CI 6-9%) overall, varying per year between 1% in 2021 to 17% in January 2022-March 2023 (95% CI 11-23%). Also in adults, SARS-CoV-2 explained only a small fraction of GAS pneumonia/sepsis in 2022-March 2023 (1%, 95% CI 0-5%). The fraction of GAS pneumonia/sepsis not attributable to respiratory viruses or non-invasive GAS was 45% (95% CI 37-53%) overall during the study period, increasing to 53% (95% CI 45-60%) in 2022-2023.

**Figure 3.**
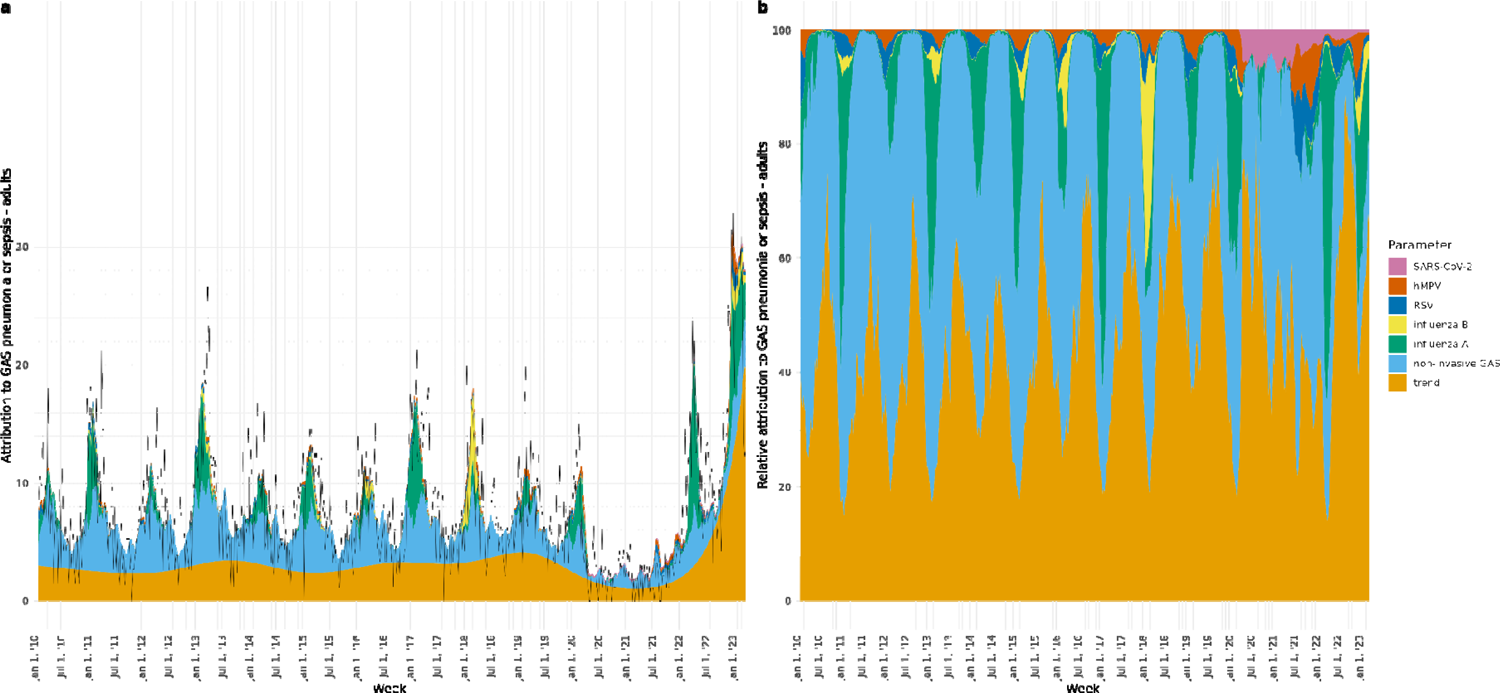
Estimated weekly absolute (A) and relative (B) attributions to parameters SARS-CoV-2, hMPV, RSV, influenza B, influenza A, non-invasive GAS and long-term time trend to GAS pneumonia or sepsis in adults, January 2010-March 2023. The black line in panel A shows the observed number of GAS pneumonia or sepsis.

**Table 3.** GAS pneumonia or sepsis in adults, attributed to respiratory virus infections, January 2010-March 2023. Absolute numbers are summarized per year and the study period, while the relative attributions are averaged over years and the study period. *: the relative attributions to SARS-CoV-2 are averaged over April-December 2020 (for the year 2020) and over April 2020-March 2023 (overall). CI: Confidence interval.

|  | Influenza A |  | Influenza B |  | RSV |  | hMPV |  | SARS- CoV-2 |  | Respiratory viruses combined |  |
| --- | --- | --- | --- | --- | --- | --- | --- | --- | --- | --- | --- | --- |
| Year | Absolute (95% CI) | Relative (95% CI) | Absolute (95% CI) | Relative (95% CI) | Absolute (95% CI) | Relative (95% CI) | Absolute (95% CI) | Relative (95% CI) | Absolute (95% CI) | Relative * (95% CI) | Absolute (95% CI) | Relative (95% CI) |
| 2010 | 23 (5-55) | 6% (1-12%) | 0 (0-1) | 0% (0-0%) | 6 (0-18) | 2% (0-5%) | 4 (0-11) | 1% (0-3%) |  |  | 34 (13-66) | 8% (3-15%) |
| 2011 | 57 (23-97) | 8% (4-11%) | 4 (1-7) | 1% (0-1%) | 8 (0-23) | 1% (0-4%) | 4 (0-12) | 1% (0-3%) |  |  | 74 (39-115) | 11% (7-15%) |
| 2012 | 23 (5-55) | 4% (1-9%) | 1 (0-1) | 0% (0-0%) | 7 (0-18) | 2% (0-4%) | 4 (0-10) | 1% (0-2%) |  |  | 34 (12-66) | 7% (3-12%) |
| 2013 | 74 (31-123) | 10% (5-14%) | 8 (2-15) | 1% (0-2%) | 6 (0-17) | 1% (0-3%) | 5 (0-14) | 1% (0-3%) |  |  | 94 (50-143) | 13% (8-17%) |
| 2014 | 39 (11-76) | 8% (3-14%) | 0 (0-1) | 0% (0-0%) | 5 (0-13) | 1% (0-3%) | 4 (0-12) | 1% (0-3%) |  |  | 49 (20-84) | 10% (5-16%) |
| 2015 | 45 (16-81) | 8% (3-12%) | 6 (1-11) | 1% (0-2%) | 6 (0-15) | 1% (0-3%) | 7 (0-19) | 1% (0-4%) |  |  | 63 (34-99) | 11% (7-15%) |
| 2016 | 27 (8-55) | 5% (2-10%) | 11 (2-21) | 2% (0-4%) | 6 (0-16) | 1% (0-3%) | 6 (0-17) | 1% (0-4%) |  |  | 50 (28-79) | 10% (6-15%) |
| 2017 | 84 (47-124) | 11% (7-14%) | 3 (0-5) | 1% (0-1%) | 5 (0-15) | 1% (0-2%) | 7 (0-20) | 1% (0-4%) |  |  | 99 (62-139) | 13% (10-17%) |
| 2018 | 18 (1-55) | 3% (0-8%) | 50 (10-93) | 7% (2-12%) | 7 (0-19) | 1% (0-4%) | 10 (0-27) | 2% (0-5%) |  |  | 85 (49-127) | 13% (9-17%) |
| 2019 | 28 (6-60) | 6% (2-11%) | 0 (0-1) | 0% (0-0%) | 6 (0-16) | 1% (0-3%) | 9 (0-25) | 2% (0-6%) |  |  | 43 (18-75) | 9% (4-14%) |
| 2020 | 40 (14-69) | 9% (4-14%) | 1 (0-2) | 0% (0-1%) | 4 (0-12) | 1% (0-3%) | 7 (0-18) | 2% (0-5%) | 5 (0-16) | 5% (0-17%) | 56 (30-87) | 16% (9-26%) |
| 2021 | 3 (2-4) | 1% (1-2%) | 0 (0-0) | 0% (0-0%) | 9 (0-26) | 5% (0-12%) | 9 (0-26) | 4% (0-12%) | 7 (0-23) | 5% (0-15%) | 28 (10-51) | 15% (6-27%) |
| 2022- Mar 2023 | 209 (116-317) | 17% (11-23%) | 25 (5-46) | 1% (0-3%) | 25 (1-68) | 3% (0-8%) | 23 (1-64) | 2% (0-6%) | 9 (0-30) | 1% (0-5%) | 291 (198-398) | 25% (18-31%) |
| Overall | 670 (500-857) | 8% (6-9%) | 110 (21-204) | 1% (0-2%) | 101 (4-275) | 2% (0-4%) | 100 (2-276) | 2% (0-4%) | 21 (1-69) | 3% (0-11%) | 1001 (804-1208) | 13% (10-16%) |

## Discussion

Our study aimed to quantify the proportions of GAS SSTI in children and GAS pneumonia/sepsis in children and adults, attributable to the circulation of several common viral infections and non-invasive GAS in the Netherlands from January 2010 through March 2023. Our model results provide a quantitative estimation of the proportion of iGAS cases that could be explained by re-emergence of predisposing viral infections during the period of excessive iGAS incidence in 2022-2023. We estimated on average 50% (95%CI 36-64%) of GAS SSTI in children to be attributable to varicella during the whole study period, but this attributable fraction was 34% (95% CI 24-43%) in the outbreak period January 2022-March 2023. Averaged over the study period, 30% (95%CI 22-39%) of GAS pneumonia/sepsis cases in children were attributable to influenza A, influenza B, RSV, hMPV and SARS-CoV-2 combined. This proportion was 37% (95%CI 23-53%) in 2022-March 2023. In adults, the attributable fraction of these respiratory viruses averaged over the study period was 13% (95%CI 10-16%), and 25% in 2022-March 2023 (95%CI 18-31%).

The attributable proportion of GAS SSTI in children to varicella per year was consistently high around 50% up to and including 2021. The high varicella incidence in 2022 after two years of very low incidence provided a likely explanation for the high iGAS SSTI incidence observed among children in the Netherlands in early 2022 [1], with 10 notifications of necrotizing fasciitis compared to 1 notification in 2016-2021 combined. However, the average attributable fraction was lower in 2022-March 2023 (34%) compared to previous years. Early 2023, the varicella incidence was very low but the number of GAS SSTI remained elevated above prepandemic levels. Therefore, varicella does not provide a sufficient explanation for the increase in GAS SSTI observed among children in 2022-2023.

GAS pneumonia/sepsis in children and adults was partly attributable to respiratory virus infections, in line with clinical observations of GAS pneumonia and pleural empyema with viral co-infections [11, 20]. While the proportion attributable to respiratory virus infections was quite high in 2022-2023 (37% for children, 25% for adults), the majority of GAS pneumonia/sepsis could not be attributed to either respiratory viruses or non-invasive GAS infections. SARS-CoV-2 does not seem to be an important risk factor for GAS pneumonia/sepsis in either children or adults; the relatively high attributable fraction of GAS pneumonia/sepsis in children averaged over the study period is mainly influenced by large fractions in 2020 and 2021, which are based on low case numbers.

The large proportion of GAS SSTI and pneumonia/sepsis, both in children and adults, that remains unexplained by respiratory viruses or non-invasive GAS in 2022-2023, and attributed to the large scale time trend parameter, suggests an - as yet-unmeasured risk factor increasing in the end of the study period. Possibly, this relates to virulence changes in circulating GAS. In 2019, Lynskey et al first described a new *emm*1 lineage among GAS, called M1*_UK_*, showing increased toxin production in vitro and replacing the previous *emm*1 strain of GAS [21]. In 2019, M1*_UK_* comprised 64% of *emm*1 iGAS isolates in the Netherlands [22]. At the end of 2022, the proportion of *emm*1 among iGAS isolates increased rapidly from 32% in November to 68% in December [1], and stayed >50% up to the end of our study period. Preliminary data (Rümke et al, unpublished) shows that M1*_UK_* has near completely replaced M1*_global_* strains in the Netherlands, with 86% M1UK variants among 267 *emm*1.0 isolates from 2022-2023. Further evolution of M1*_UK_*, with acquisition of additional virulence genes, has been described [23], as well as another *emm*1.0 variant emerging in Denmark [24]. Increased virulence of GAS due to emerging variants may well have contributed to the increase in iGAS seen in 2022-2023, and account for (a part of) the unexplained proportions as seen in Figures 1-3 (“trend” parameter). Alternatively, increased susceptibility in the population due to years of reduced exposure (‘immunity debt’) has been proposed as an explanation for the high iGAS incidence. However, such a generally increased susceptibility would likely also be reflected by high numbers of non-invasive GAS syndrome consultations, which we did not observe.

Important limitations of our study include the proxies used for GAS SSTI and pneumonia/sepsis. GAS cultures from skin, pus and wound are not specific for invasive GAS and may include cultures from patients with noninvasive disease such as impetigo or abscesses. Conversely, for some patients with severe SSTI GAS is confirmed by blood culture only and is therefore not included in the SSTI numbers. Likewise, the blood and lower airway cultures are not specific for GAS pneumonia/sepsis. Therefore, it is difficult to interpret our results in absolute numbers of iGAS infections or disease burden attributable to the pathogens included. Also, as the catchment populations of the participating laboratories are unknown and can vary over time, our adjustment for the number of laboratories contributing to the data likely does not fully capture variation in coverage. A limitation of the virus detection data for influenza A and B, RSV and hMPV is that these cannot be stratified into age groups and detections may be influenced by changes in testing policies, especially during the pandemic, affecting the time series.

Another limitation of our study is the implicit assumption that the models were correctly specified. This assumption could not be fully tested. Results could change if we would have included other pathogens, or excluded some of the pathogens we have calculated the attribution for. This means that the model estimates should be interpreted with care. Also, in time series modelling, harmonic terms are often included to allow for seasonal baseline fluctuations in the outcome variable. As we included data on consultations for non-invasive GAS syndromes, which incorporates the winter seasonality of GAS infections in general, we chose not to include harmonic terms. Had we done so, possibly the proportions attributed to e.g. varicella and influenza would have been lower. On the other hand, the choice to model the long-term time trend using a penalized spline allowed for the absorption of unaccounted effects, thereby mitigating model misspecification. The flexibility of the spline term enables it to capture variations in the data that might not be adequately addressed by the predefined model structure. This absorption of unexplained variability into the spline term enhances the model’s ability to fit the data accurately and reduces the potential for misrepresenting relationships. Had we used a less flexible time trend, such as a linear term or constant intercept, likely the attribution to mainly influenza A would have been larger, as the coefficient for this pathogen was allowed to vary per season.

A final limitation is the fact that this is an ecological study of temporal associations. The associations we reported do not necessarily reflect causal effects, although such causality has been established by other studies. Effects of influenza A infection on iGAS infection and severity has been shown in murine studies [9, 25]. Varicella is a well-known risk factor for iGAS based on epidemiological studies [7, 8].

A strength of our study is the inclusion of lags 0-4 weeks of all pathogens on the outcomes, by using a distributed lag model. Unlike fixed lag models, which can be biased if lag choice misaligns with the true data generating process, distributed lag models mitigate this by considering a range of lags. The distributed lag model provides a data-driven way to identify and capture appropriate delayed effects [17]. Another strength is the length of the study period. This allowed for robust estimation of pre-pandemic associations for comparison to the 2022-2023 situation. While associations between iGAS and both influenza and varicella have been described before, our study further quantifies the associations between iGAS and RSV, hMPV, and SARS-CoV-2 and their relative importance.

In the Netherlands, vaccination against varicella is not included in the national childhood immunization programme. Seasonal influenza vaccination is offered to adults aged 60 years and older, and to persons of any age with certain medical conditions. Uptake of the seasonal influenza vaccine among eligible persons was estimated at 58% in 2021 [26]. Our results suggest that introducing varicella vaccination or expanding influenza vaccination eligibility could have an effect on iGAS disease burden. Indeed, studies from Israel and Canada have shown a decrease in iGAS incidence after introduction of varicella vaccination [27, 28].

In conclusion, our results point to relevant attributable fractions of GAS SSTI among children to varicella and of GAS pneumonia/sepsis to influenza A in adults and in children aged 0-5 years, and additional relevant attributable fractions of GAS pneumonia/sepsis in children to RSV and hMPV. However, the re-emergence of these viruses does not provide a sufficient explanation for the surge in iGAS infections in 2022-2023 in either children or adults in the Netherlands. Other factors increasing iGAS risk, such as microbial virulence factor acquisition, may be at play.

## Ethical statement

The Centre for Clinical Expertise at the RIVM verified whether this work complied with either the specific conditions as stated in article one of the Dutch law for Medical Research Involving Human Subjects (WMO) or with the EU Clinical Trial Directive (2001/20/EC), and was of the opinion that the research did not fulfill these conditions. Therefore, no formal ethical approval was required.

## Author contributions

Conceptualization: BdG, JvdK, LvA, AS, MtW. Data analysis: JvdK, BdG. First draft of manuscript: BdG, JvdK. Critical revision of manuscript: HEdM, NMvS, ISIS-AR study group, MH, MtW, JvdK, LvA, AS, BdG. All authors approved the final manuscript.

## Data Availability

All data and code produced in the present study are available at https://github.com/kassteele/Papers/DeGier_iGAS_attribution

## Acknowledgements

We thank Rony Zoetigheid and Britt de Wit (Center for Infectious Disease Control, RIVM) for preparing timeseries datasets. We thank the Nivel Primary Care Database team, participating general practitioners and their patients. We also thank the Dutch Working Group on Clinical Virology from the Dutch Society for Clinical Microbiology (NVMM) and all participating laboratories for providing the virological data from the weekly Sentinel Surveillance system.

## Members of the ISIS-AR study group

J.W.T. Cohen Stuart, Noordwest Ziekenhuisgroep, Department of Medical Microbiology, Alkmaar D.C. Melles, Meander Medical Center, Department of Medical Microbiology, Amersfoort A. K. van Dijk, Amsterdam UMC, University of Amsterdam, Department of Medical Microbiology and Infection Prevention, Amsterdam Infection and Immunity Institute, Amsterdam A. Alzubaidy, Atalmedial, Department of Medical Microbiology, Amsterdam B. M. Scholing, OLVG Lab BV, Department of Medical Microbiology, Amsterdam S.D. Kuil, Public Health Service, Public Health Laboratory, Amsterdam G.J. Blaauw, Gelre Hospitals, Department of Medical Microbiology and Infection prevention, Apeldoorn ISIS-AR Projectteam: W. Altorf - van der Kuil, S.M. Bierman, S.C. de Greeff, S.R. Groenendijk, R. Hertroys, J.C.M Monen, D.W. Notermans, J. Polman, W.J. van den Reek, C. Schneeberger-van der Linden, A.F. Schoffelen, F. Velthuis, C.C.H. Wielders, B.J. de Wit, R.E. Zoetigheid, Centre for Infectious Disease Control (CIb), National Institute for Public Health and the Environment (RIVM), Bilthoven, The Netherlands A. W. van den Bijllaardt, Microvida Amphia, Laboratory for Microbiology and Infection Control, Breda E.M. Kraan, IJsselland hospital, Department of Medical Microbiology, Capelle a/d Ijssel M.B. Haeseker, Reinier de Graaf Group, Department of Medical Microbiology, Delft J.M. da Silva, Deventer Hospital, Department of Medical Microbiology, Deventer A. E. de Jong, Slingeland Hospital, Department of Medical Microbiology, Doetinchem B. Maraha, Albert Schweitzer Hospital, Department of Medical Microbiology, Dordrecht A.J. van Griethuysen, Gelderse Vallei Hospital, Department of Medical Microbiology, Ede B.B. Wintermans, Admiraal De Ruyter Hospital, Department of Medical Microbiology, Goes M.J.C.A. van Trijp, Groene Hart Hospital, Department of Medical Microbiology and Infection Prevention, Gouda A.E. Muller, Haaglanden MC, Department of Medical Microbiology, ‘s-Gravenhage M. Wong, Haga Hospital, Department of Medical Microbiology, ‘s-Gravenhage A. Ott, Certe, Medical Microbiology Groningen|Drenthe, Groningen I. E. Bathoorn, University of Groningen, University Medical Center, Department of Medical Microbiology, Groningen II. M. Lokate, University of Groningen, University Medical Center, Department of Medical Microbiology, Groningen III. J. Sinnige, Regional Public Health Laboratory Haarlem, Haarlem D.C. Melles, St Jansdal Hospital, Department of Medical Microbiology, Harderwijk N. Plantinga, Laboratory of Medical Microbiology and Public Health, Hengelo N.H. Renders, Jeroen Bosch Hospital, Department of Medical Microbiology and Infection Control, ‘s-Hertogenbosch J.W. Dorigo-Zetsma, CBSL, Tergooi MC, Department of Medical Microbiology, Hilversum L.J. Bakker, CBSL, Tergooi MC, Department of Medical Microbiology, Hilversum I. K. Waar, Certe, Medical Microbiology Friesland|NOP, Leeuwarden M.T. van der Beek, Leiden University Medical Center, Department of Medical Microbiology, Leiden M.A. Leversteijn-van Hall, Eurofins Clinical Diagnostics, Department of Medical Microbiology, Leiden-Leiderdorp S.P. van Mens, Maastricht University Medical Centre, Department of Medical Microbiology, Maastricht I. E. Schaftenaar, St Antonius Hospital, Department of Medical Microbiology and Immunology, Nieuwegein M.H. Nabuurs-Franssen, Canisius Wilhelmina Hospital, Department of Medical Microbiology and Infectious Diseases, Nijmegen I. Maat, Radboud University Medical Center, Department of Medical Microbiology, Nijmegen P.D.J. Sturm, Laurentius Hospital, Roermond B.M.W. Diederen, Bravis Hospital, Department of Medical Microbiology, Roosendaal L.G.M. Bode, Erasmus University Medical Center, Department of Medical Microbiology and Infectious Diseases, Rotterdam D.S.Y. Ong, Franciscus Gasthuis and Vlietland, Department of Medical Microbiology and Infection Control, Rotterdam A. M. van Rijn, Ikazia Hospital, Department of Medical Microbiology, Rotterdam S. Dinant, Maasstad Hospital, Department of Medical Microbiology, Rotterdam M. den Reijer, Star-SHL, Rotterdam D.W. van Dam, Zuyderland Medical Centre, Department of Medical Microbiology and Infection Control, Sittard-Geleen E.I.G.B. de Brauwer, Zuyderland Medical Centre, Department of Medical Microbiology and Infection Control, Sittard-Geleen R.G. Bentvelsen, Microvida ZorgSaam, Department of Medical Microbiology, Terneuzen A.G.M. Buiting, St. Elisabeth Hospital, Department of Medical Microbiology, Tilburg A.L.M. Vlek, Diakonessenhuis, Department of Medical Microbiology and Immunology, Utrecht M. de Graaf, Saltro Diagnostic Centre, Department of Medical Microbiology, Utrecht A. Troelstra, University Medical Center Utrecht, Department of Medical Microbiology, Utrecht A.R. Jansz, Eurofins-PAMM, Department of Medical Microbiology, Veldhoven M.P.A. van Meer, Rijnstate Hospital, Laboratory for Medical Microbiology and Immunology, Velp J. de Vries, VieCuri Medical Center, Department of Medical Microbiology, Venlo J. D. Machiels, Isala Hospital, Laboratory of Medical Microbiology and Infectious Diseases, Zwolle

---

**Figure S1.**
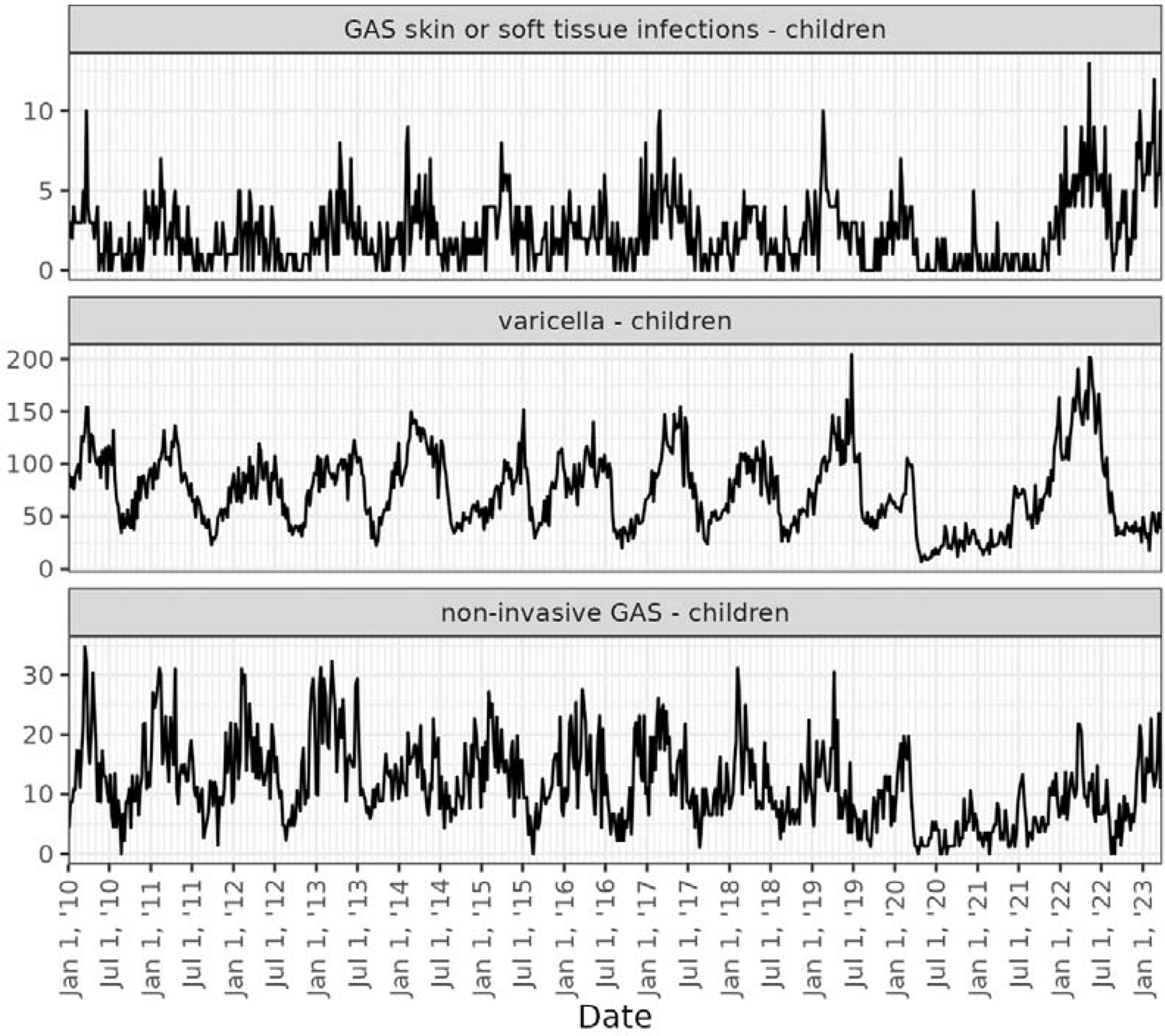
Observed weekly numbers of GAS skin and soft tissue infections in children aged 0-5 years, varicella consultations and non-invasive GAS infection consultations per 100.000 children age 0-4, 1 January 2010-March 2023.

**Figure S2.**
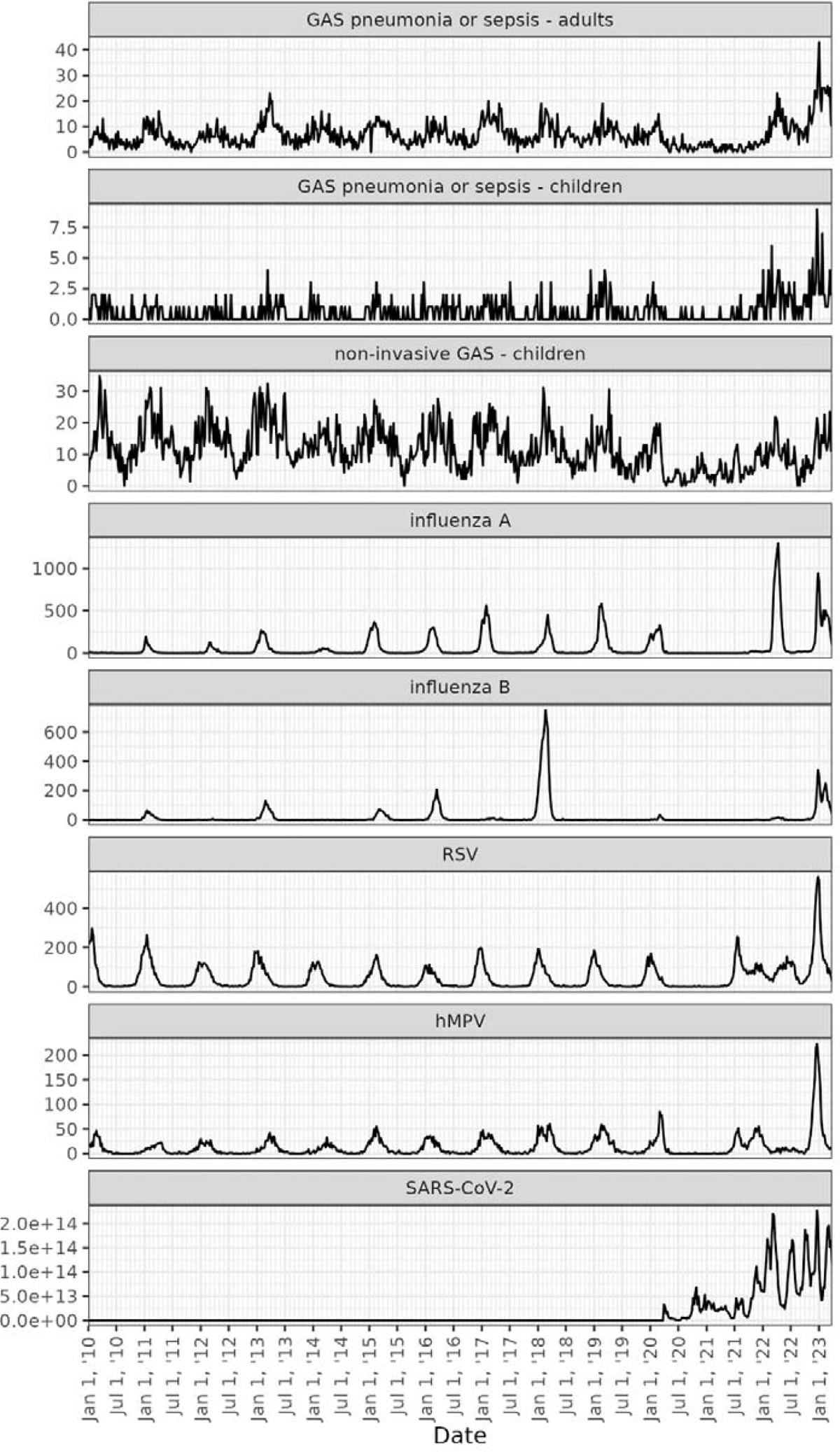
Observed weekly numbers of GAS pneumonia or sepsis in adults age 18 and older and in children aged 0-5 years, and detections of influenza A, influenza B, respiratory syncytial virus (RSV), human metapneumovirus (hMPV), and average sewage RNA load of severe acute respiratory syndrome coronavirus 2 (SARS-CoV-2) per 100.000 inhabitants, 1 January 2010-March 2023.

**Figure S3.**
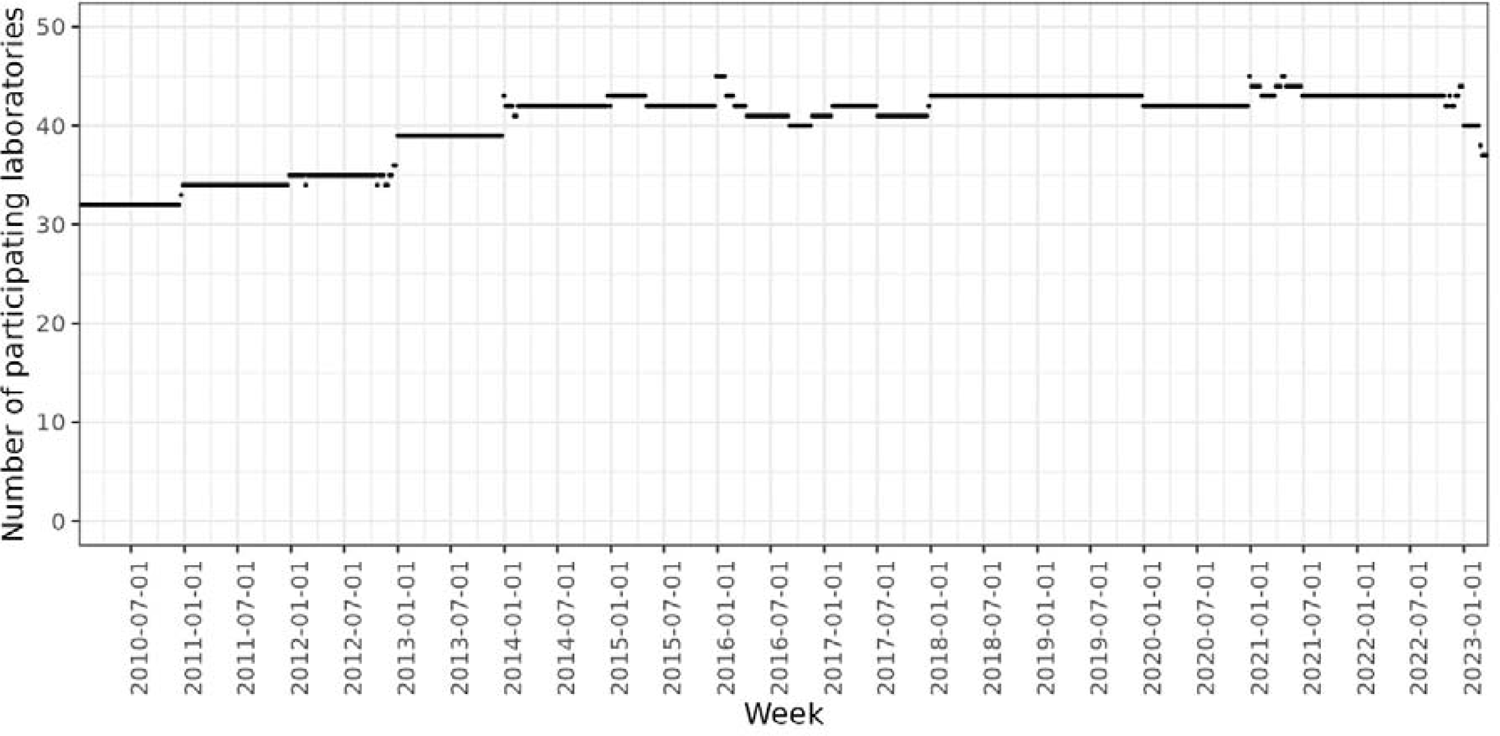
Number of laboratories contributing data to ISIS-AR per week during the study period (January 2010-March 2023).

## References

1. de Gier B, Marchal N, de Beer-Schuurman I, Te Wierik M, Hooiveld M, Group I-AS, et al. Increase in invasive group A streptococcal (Streptococcus pyogenes) infections (iGAS) in young children in the Netherlands, 2022. Euro Surveill. 2023;28(1).

2. Increase in invasive group A streptococcal infections among children in Europe, including fatalities. [press release]. Stockholm: European Centre for Disease Prevention and Control (ECDC) 2022.

3. Guy R, Henderson KL, Coelho J, Hughes H, Mason EL, Gerver SM, et al. Increase in invasive group A streptococcal infection notifications, England, 2022. Euro Surveill. 2023;28(1).

4. Lassoued Y, Assad Z, Ouldali N, Caseris M, Mariani P, Birgy A, et al. Unexpected Increase in Invasive Group A Streptococcal Infections in Children After Respiratory Viruses Outbreak in France: A 15-Year Time-Series Analysis. Open Forum Infect Dis. 2023;10(5):ofad188.

5. Gouveia C, Bajanca-Lavado MP, Mamede R, Araujo Carvalho A, Rodrigues F, Melo-Cristino J, et al. Sustained increase of paediatric invasive Streptococcus pyogenes infections dominated by M1(UK) and diverse emm12 isolates, Portugal, September 2022 to May 2023. Euro Surveill. 2023;28(36).

6. Cobo-Vazquez E, Aguilera-Alonso D, Carrasco-Colom J, Calvo C, Saavedra-Lozano J, Ped GASnWG. Increasing incidence and severity of invasive Group A streptococcal disease in Spanish children in 2019-2022. Lancet Reg Health Eur. 2023;27:100597.

7. Laupland KB, Davies HD, Low DE, Schwartz B, Green K, McGeer A. Invasive group A streptococcal disease in children and association with varicella-zoster virus infection. Ontario Group A Streptococcal Study Group. Pediatrics. 2000;105(5):E60.

8. Imohl M, van der Linden M, Reinert RR, Ritter K. Invasive group A streptococcal disease and association with varicella in Germany, 1996-2009. FEMS Immunol Med Microbiol. 2011;62(1):101–9.

9. Herrera AL, Huber VC, Chaussee MS. The Association between Invasive Group A Streptococcal Diseases and Viral Respiratory Tract Infections. Front Microbiol. 2016;7:342.

10. de Gier B, Vlaminckx BJM, Woudt SHS, van Sorge NM, van Asten L. Associations between common respiratory viruses and invasive group A streptococcal infection: A time-series analysis. Influenza Other Respir Viruses. 2019;13(5):453–8.

11. van Kempen EB, Bruijning-Verhagen PCJ, Borensztajn D, Vermont CL, Quaak MSW, Janson JA, et al. Increase in Invasive Group a Streptococcal Infections in Children in the Netherlands, A Survey Among 7 Hospitals in 2022. Pediatr Infect Dis J. 2023;42(4):e122–e4.

12. Altorf-van der Kuil W, Schoffelen AF, de Greeff SC, Thijsen SF, Alblas HJ, Notermans DW, et al. National laboratory-based surveillance system for antimicrobial resistance: a successful tool to support the control of antimicrobial resistance in the Netherlands. Euro Surveill. 2017;22(46).

13. Nivel. Actuele Weekcijfers Aandoeningen-Surveillance. 2023 [Available from: https://www.nivel.nl/nl/nivel-zorgregistraties-eerste-lijn/cijfers-over-aandoeningen/actuele-weekcijfers-aandoeningen-surveillance.

14. van Boven M, Hetebrij WA, Swart A, Nagelkerke E, van der Beek RF, Stouten S, et al. Patterns of SARS-CoV-2 circulation revealed by a nationwide sewage surveillance programme, the Netherlands, August 2020 to February 2022. Euro Surveill. 2023;28(25).

15. Eilers PHC, Marx BD. Flexible smoothing with *B*-splines and penalties. Statistical Science. 1996;11(2):89–121, 33.

16. Scheipl F. spikeSlabGAM: Bayesian Variable Selection, Model Choice and Regularization for Generalized Additive Mixed Models in R. Journal of Statistical Software. 2011;43(14):1–24.

17. Gasparrini A. Distributed Lag Linear and Non-Linear Models in R: The Package dlnm. J Stat Softw. 2011;43(8):1–20.

18. R: A language and environment for statistical computing. R Foundation for Statistical Computing, Vienna, Austria: R Core Team 2022 [Available from: https://www.R-project.org/.

19. RStan: the R interface to Stan. R package version 2.21.8.. Stan Development Team 2023.

20. Davies PJB, Russell CD, Morgan AR, Taori SK, Lindsay D, Ure R, et al. Increase of Severe Pulmonary Infections in Adults Caused by M1(UK) Streptococcus pyogenes, Central Scotland, UK. Emerg Infect Dis. 2023;29(8):1638–42.

21. Lynskey NN, Jauneikaite E, Li HK, Zhi X, Turner CE, Mosavie M, et al. Emergence of dominant toxigenic M1T1 Streptococcus pyogenes clone during increased scarlet fever activity in England: a population-based molecular epidemiological study. Lancet Infect Dis. 2019;19(11):1209–18.

22. Rumke LW, de Gier B, Vestjens SMT, van der Ende A, van Sorge NM, Vlaminckx BJM, et al. Dominance of M1(UK) clade among Dutch M1 Streptococcus pyogenes. Lancet Infect Dis. 2020;20(5):539–40.

23. Davies MR, Keller N, Brouwer S, Jespersen MG, Cork AJ, Hayes AJ, et al. Detection of Streptococcus pyogenes M1(UK) in Australia and characterization of the mutation driving enhanced expression of superantigen SpeA. Nat Commun. 2023;14(1):1051.

24. Johannesen TB, Munkstrup C, Edslev SM, Baig S, Nielsen S, Funk T, et al. Increase in invasive group A streptococcal infections and emergence of novel, rapidly expanding sub-lineage of the virulent Streptococcus pyogenes M1 clone, Denmark, 2023. Euro Surveill. 2023;28(26).

25. Herrera AL, Potts R, Huber VC, Chaussee MS. Influenza enhances host susceptibility to non-pulmonary invasive Streptococcus pyogenes infections. Virulence. 2023;14(1):2265063.

26. Heins M, Korevaar J, Knottnerus B, Hooiveld M. Vaccine Coverage Dutch National Influenza Prevention Program 2021: brief monitor.. In: Nivel, editor. 2022.

27. Frere J, Bidet P, Tapiero B, Rallu F, Minodier P, Bonacorsi S, et al. Clinical and Microbiological Characteristics of Invasive Group A Streptococcal Infections Before and After Implementation of a Universal Varicella Vaccine Program. Clin Infect Dis. 2016;62(1):75–7.

28. Hasin O, Hazan G, Rokney A, Dayan R, Sagi O, Ben-Shimol S, et al. Invasive Group A Streptococcus Infection in Children in Southern Israel Before and After the Introduction of Varicella Vaccine. J Pediatric Infect Dis Soc. 2020;9(2):236–9.

